# Development and Validation of Postoperative Venous Thromboembolism risk prediction model

**DOI:** 10.1101/2020.06.21.20136432

**Authors:** Sang H. Woo, Ruben Rhoades, Lily Ackermann, Scott W. Cowan, Jillian Zavodnick, Gregary D. Marhefka

**Author notes:** **Correspondence:** Sang Hoon Woo, M.D., Division of Hospital Medicine, Department of Medicine, Thomas Jefferson University Suite 701, 833 Chestnut St., Philadelphia, PA 19107. No funding was obtained for this manuscript. The content of this manuscript was not presented at any prior conferences.

## Abstract

**Background:** VTE is a serious postoperative complication after surgery with resultant higher morbidity and mortality. Despite years of experience with current risk models, rates continue to be high and more information is needed on individual patient risk in the prophylaxis era.

**Research Questions:** Can we assess the individualized risk of postoperative venous thromboembolism (VTE) for broad categories of surgery?

**Methods:** This study was performed using data from the American College of Surgeons National Surgical Quality Improvement Program (ACS-NSQIP) Database. Patient data (n=2,875,190) from 2015-2017 were used for study analysis. Eight predictors were selected for the model: age, preoperative platelet count≥450 (×10^9^/L), disseminated cancer, corticosteroid use, serum albumin ≤2.5 g/dL, preoperative sepsis, hospital length of stay and surgery type. The second model included 7 predictors without hospital length of stay. A predictive model was trained using ACS-NSQIP data from 2015-2016 (n=1,859,227) and tested using data from 2017 (n= 1,015,963). Primary outcomes are postoperative 30-day VTE, including deep vein thrombosis (DVT) and/or pulmonary embolism (PE).

**Results:** VTE occurred in 23,249 patients (0.81%) and 49.9% of VTE occurred after discharge from index hospitalization. The risk prediction model had high AUC (area under the receiver operating characteristic curve) for postoperative VTE of 0.78 (training cohort) and 0.78 (test cohort).

**Interpretation:** This clinical prediction model is a validated, practical and easy-to-use tool to identify surgical patients at the highest risk of postoperative VTE and provide an individualized assessment of risk based on clinical factors and type of surgery. This prediction model may be used as a tool to assess individualized risk of postoperative VTE and promote broader discussion and awareness of the VTE risk during the perioperative period.

## INTRODUCTION

Large numbers of surgical and medical patients are at risk of venous thromboembolism. ^1,2^ Postoperative deep venous thrombosis (DVT) and pulmonary embolism (PE) are major surgical complications with high mortality and morbidity. ^3–5^ To reduce the risk of these complications, the American College of Chest Physicians (ACCP) published guidelines recommending the use of pharmacologic and/or mechanical prophylaxis according to risk scores.^6^ Despite the use of various thromboprophylaxis methods, the incidence of postoperative venous thromboembolism remains high. ^7^ Accurate assessment of a patient’s VTE risk helps to foster awareness and discussion with the patient and the surgeon during the perioperative period while increasing compliance with evolving prophylaxis guidelines.

Various qualitative and quantitative risk assessment models have been developed and are in use. ^8–11^ However, currently available venous thromboembolism risk assessment models have limitations. The modified Caprini Risk Assessment model and the Rogers score have been frequently used, ^8^ but these models were developed from a limited number of surgical procedures and before current pharmacologic prophylaxis methods, such as low molecular weight heparin (LMWH) and direct oral anticoagulants (DOACs), were widely utilized. Thus, a need exists for a risk assessment model developed from a contemporary patient population, that is applicable to -- and validated in -- broader types of surgery.

Pharmacologic methods for postoperative VTE prophylaxis are highly variable, including aspirin, unfractionated heparin, LMWH, or DOACs. Current guidelines allow the use of different anticoagulation approaches but provide limited guidance for which patients warrant which pharmacologic method. More targeted, individualized preventive measures for patients at particularly high risk may help further lower rates of VTE by guiding clinicians toward more aggressive prophylaxis. ^12 6^ In order to accurately assess an individual’s risk, development of a quantitative risk assessment of VTE is necessary that can be applied to broad types of surgery, which may necessitate different types of DVT prophylaxis.

This analysis was performed using the American College of Surgeons National Surgical Quality Improvement Program (ACS-NSQIP) database.

## METHODS

### Study design, population

The ACS-NSQIP database is a prospective database of patients undergoing surgery at more than 500 US and international, academic and community hospitals. Data including preoperative demographics, risk factors, and 30-day postoperative outcomes are obtained by trained surgical clinical reviewers at each participating hospital for submission to the database. To develop this model, we analyzed 2,875,190 patients in the ACS-NSQIP database from 2015 to 2017.

Risk factors included in our analysis were age, sex, smoking, history of congestive heart failure, history of chronic obstructive pulmonary disease, diabetes, preoperative sepsis, use of anti-hypertensive medications, chronic corticosteroid use, disseminated cancer, dialysis, preoperative renal failure, admission origin, body mass index, admission type (inpatient, outpatient status), American Society of Anesthesiologists’ (ASA) class, preoperative functional status, and the type of surgery. Preoperative laboratory tests in the analysis include serum sodium, creatinine, white blood cell count, hematocrit, and platelet count.

### Outcome

Occurrence of VTE, including DVT and/or PE, within 30 days of surgery was the primary outcome.

### Data Analysis

The predictive model was trained using data from years 2015 and 2106 (N= 1,859,227) and validated using data from 2017 (N=1,015,963). Clinical predictors to be included in the predictive model were selected using backwards elimination, clinical relevance and related literature. Predictors considered for the model were selected without knowing the outcome based on the literature. Multivariate logistic regression analysis was performed to obtain covariate coefficient, standard error, and adjusted odd’s ratio. Missing lab values (hematocrit, platelet, albumin, white blood cell count) were recorded as a separate category. Patient demographic factors were compared using Chi-squared tests for categorical variables and Wilcoxon for continuous variables.

AUC(Area Under the Curve) was used to evaluate the model performance. We used GridSearchCV package with 5 fold cross validation to determine the best model parameters. Statistical analysis was done using Python (version 3.6.6), Statsmodels (version 0.9.0), and r programming (RStudio version 1.1.463). Scikit-learn package was used for machine learning modeling and web application. The Thomas Jefferson University Institutional Review Board approved this study and granted a waiver of informed consent from study participants.

## Results

### Patient population

**Table 1** shows demographic characteristics of study participant cohorts. VTE occurrence rate was 0.82 % (n=15,159) in 2015 to 2016. Patients who developed VTE were older (63.4 vs 56.5 years, p<0.001). Patients with postoperative VTE had higher prevalence of disseminated cancer (8.75% vs 2.22%, p<0.001), insulin dependent diabetes (7.64% vs 5.78%, p<0.001), dialysis (2.03% v 1.31%, p<0.001), and COPD (7.96% vs 4.45%, p<0.001). **Figure 1** shows 30-day mortality and readmission rates associated with the occurrences of VTE. Patient who developed VTE had higher rates of death (5.9% vs 0.9%), and 30 day readmission rates (37.5% vs 5.0%)

**Table 1.**
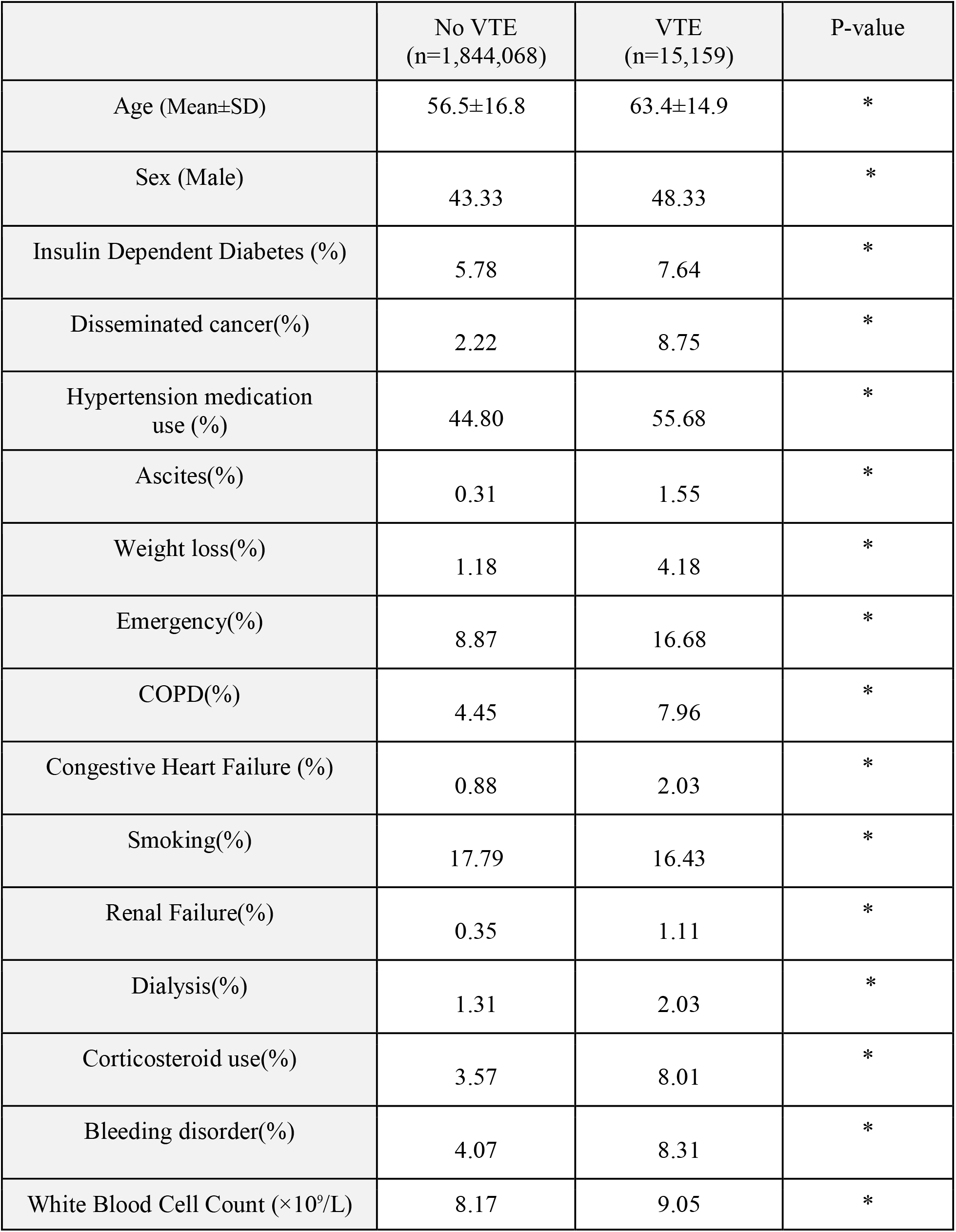
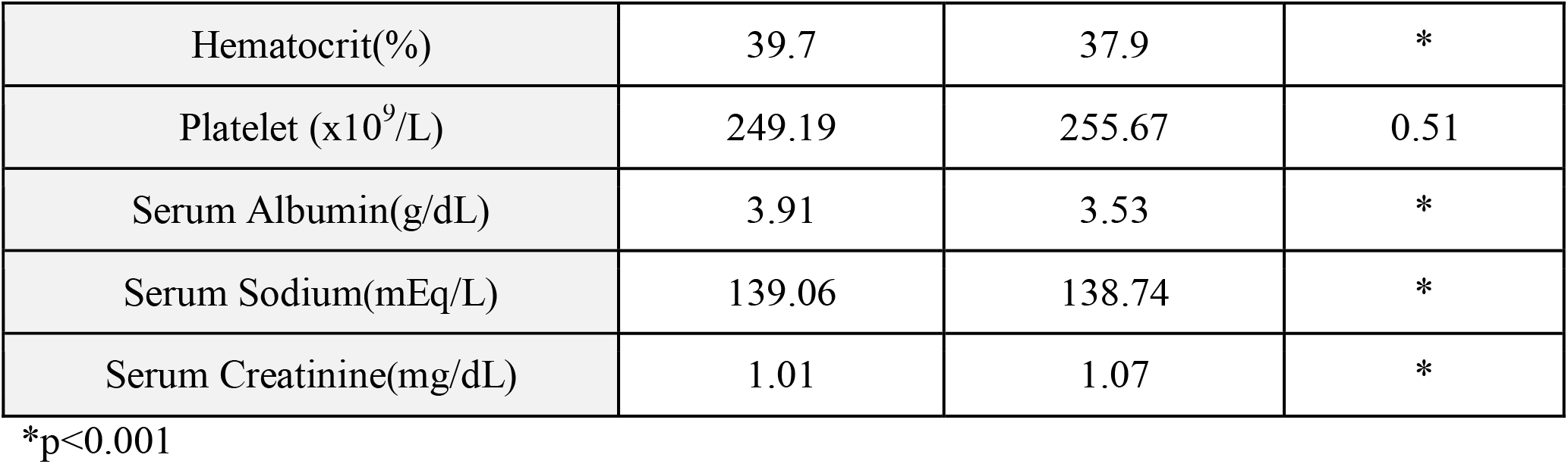
Characteristics of the patients (patients from 2015 to 2016)

**Figure 1.**
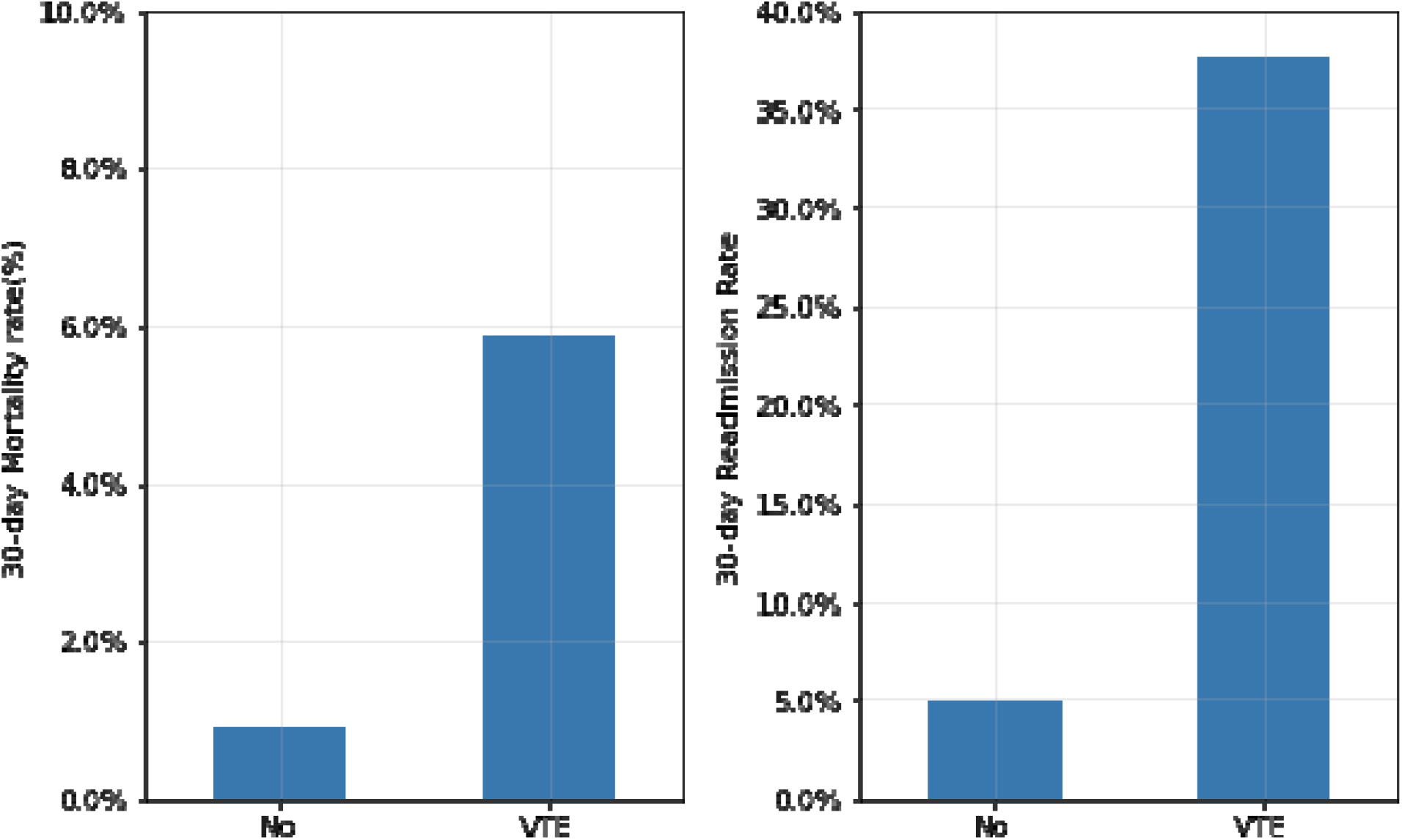
30-day postoperative mortality and readmission rates with venous thromboembolism.

### Age, Platelet Count, Serum Albumin and Incidence of VTE

**Figure 2** illustrates the unadjusted rates of postoperative VTE according to age, platelet count, and serum albumin level from years 2015 to 2017. Both thrombocytopenia and thrombocytosis were associated with increased unadjusted postoperative VTE rate. Patients with thrombocytosis with platelets of ≥ 450 (×10^9^/L) had a VTE rate of 2.2% compared with 0.9% for patients with platelets of < 450(×10^9^/L). Older age was associated with higher incidence of VTE. Patients age 71-80 years had a VTE rate of 1.2%, compared with 0.71% for age 46-60 years.

**Figure 2.**
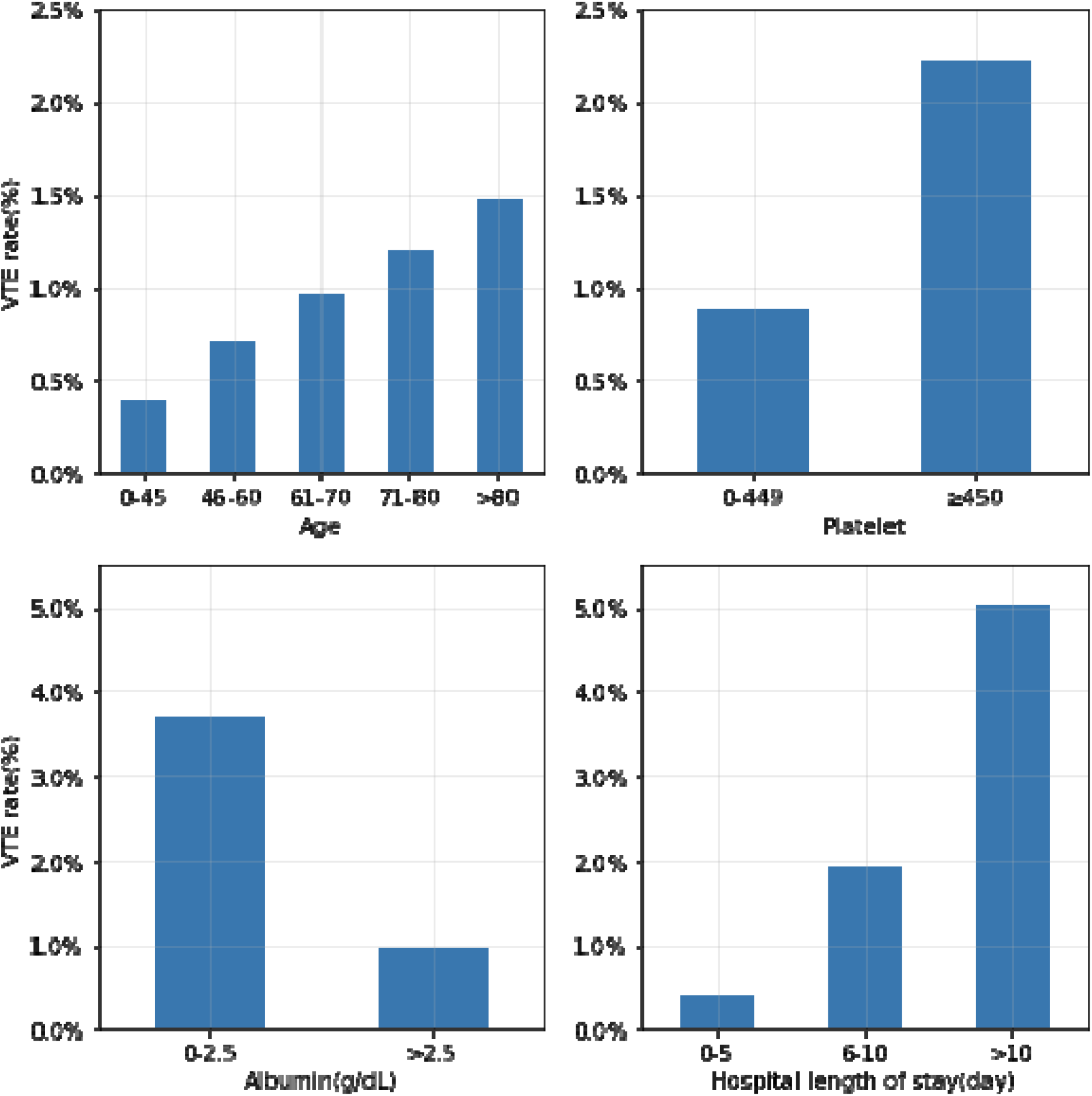
Age, Platelet count, Serum Albumin, Length of Stay and unadjusted VTE rates. Patients from 2015 to 2017

A low serum albumin level was associated with a higher incidence of VTE. Those with a serum albumin ≤ 2.5 g/dL had a VTE incidence of 3.71%, compared with 1.0% for those with albumin >2.5 g/dL.

### Surgery type and VTE

30-day postoperative VTE rates vary significantly among different types of surgery. **Figure 3** shows VTE rates according to type of surgery from 2015 to 2016 cohorts. Compared with rates of <2% for other surgery types, surgeries involving the liver, pancreas, or spleen (3.08%), brain (2.84%), and vascular-aorta (2.36%) were associated with higher VTE rates. Among orthopedic surgeries, lower extremity and pelvic surgeries were associated with higher VTE rates compared to upper extremity or shoulder surgery (1.10% vs 0.33%).

**Figure 3.**
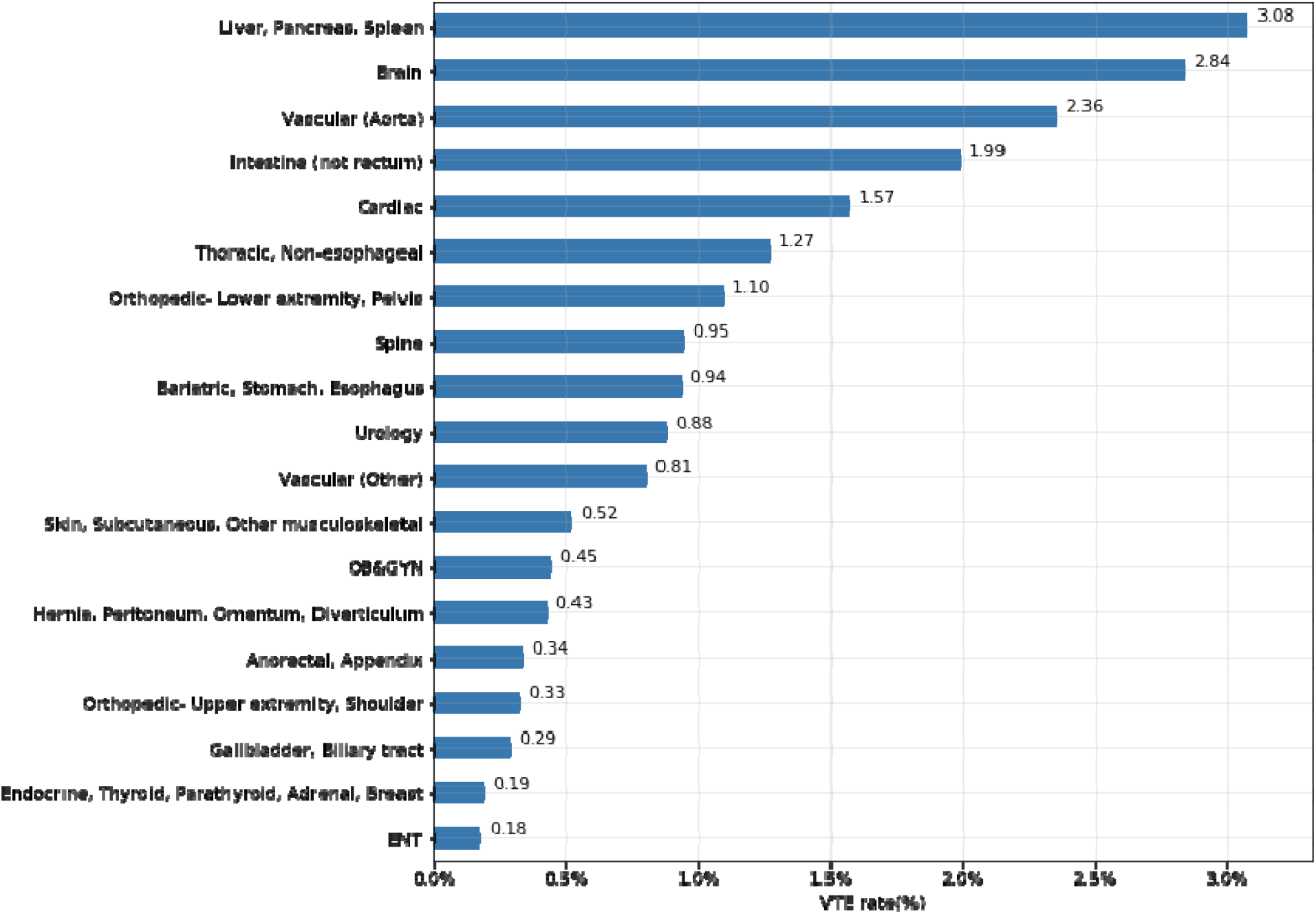
VTE rates (%) according to type of surgery. (patients from 2015-16)

49.91% of DVT and 49.93% of PE complications occurred after discharge from index hospitalization. **eFigure V** in the **Supplement** shows the average day from operation until DVT complication according to surgery type.

### Prediction and validation of the predictive model

**Table 2** shows eight predictors selected for the VTE risk assessment model. Age, disseminated cancer, hypoalbuminemia (albumin ≤2.5 g/dL), corticosteroid use and hospital length of stay had significant association with 30-day postoperative DVT occurrence. Patients with low serum albumin (albumin ≤2.5g/dL) had a high adjusted odds ratio (OR=1.17, P<0.001). Thrombocytosis (platelet ≥450×10^9^/L) also had a high adjusted odds ratio (OR= 1.38, P<0.001). As shown in **eFigure II** in the **Supplement**, the model has excellent calibration. The VTE prediction model has excellent model performance (training cohort AUC= 0.78, validation cohort AUC=0.78). Brier score was 0.008. 5-fold cross validation method for the training cohort showed mean AUC of 0.79.

**Table 2.** Adjusted odds ratio (OR) of VTE predictors.

|  | Adjusted odds ratio | coefficient | SE | P-Value |
| --- | --- | --- | --- | --- |
| Intercept |  | -6.954 | 0.146 | <0.001 |
| Age | 1.02 (1.01-1.02) | 0.015 | 0.001 | <0.001 |
| Steroid use | 1.44 (1.35-1.53) | 0.364 | 0.032 | <0.001 |
| Disseminated cancer | 1.93 (1.81-2.05) | 0.655 | 0.031 | <0.001 |
| Albumin ≤2.5 g/dL | 1.17 (1.09-1.25) | 0.154 | 0.035 | <0.001 |
| Sepsis (reference =none) |  |  |  |  |
| SIRS | 1.57 (1.46-1.68) | 0.449 | 0.036 | <0.001 |
| Sepsis | 1.65 (1.53-1.78) | 0.499 | 0.038 | <0.001 |
| Septic Shock | 2 (1.79-2.24) | 0.694 | 0.058 | <0.001 |
| Platelet (x10<sup>9</sup>/L)<br>(reference=0-449) |  |  |  |  |
| ≥450 | 1.38 (1.27-1.5) | 0.322 | 0.041 | <0.001 |
| Surgery type (reference =ENT) |  |  |  |  |
| anorectal, appendix | 1.29 (0.96-1.74) | 0.253 | 0.152 | 0.097 |
| bariatric, stomach, esophagus | 3.34 (2.51-4.46) | 1.207 | 0.147 | <0.001 |
| brain | 6.83 (5.1-9.14) | 1.921 | 0.149 | <0.001 |
| cardiac | 3.11 (2.22-4.34) | 1.133 | 0.171 | <0.001 |
| endocrine, thyroid, parathyroid,<br>adrenal, breast | 0.92 (0.69-1.25) | -0.080 | 0.152 | 0.602 |
| gallbladder, biliary tract | 1.14 (0.85-1.54) | 0.132 | 0.153 | 0.388 |
| hernia, peritoneum, omentum,<br>diverticulum | 1.74 (1.31-2.32) | 0.556 | 0.146 | <0.001 |
| intestine (not rectum) | 3.8 (2.86-5.03) | 1.334 | 0.144 | <0.001 |
| liver, pancreas, spleen | 6.31 (4.73-8.41) | 1.842 | 0.147 | <0.001 |
| obstetrics, gynecology | 2.09 (1.57-2.8) | 0.738 | 0.148 | <0.001 |
| orthopedic, lower extremity, pelvis | 3.3 (2.49-4.37) | 1.192 | 0.144 | <0.001 |
| orthopedic upper extremity,<br>shoulder | 1.51 (1.1-2.09) | 0.414 | 0.164 | 0.012 |
| skin, subcutaneous, other<br>musculoskeletal | 2.07 (1.55-2.76) | 0.726 | 0.147 | <0.001 |
| spine | 3.54 (2.66-4.71) | 1.264 | 0.146 | <0.001 |
| thoracic, non-esophageal | 2.94 (2.16-3.99) | 1.078 | 0.156 | <0.001 |
| urology | 3.08 (2.31-4.1) | 1.124 | 0.146 | <0.001 |
| aorta | 4.51 (3.19-6.38) | 1.506 | 0.177 | <0.001 |
| Vascular | 2.28 (1.71-3.04) | 0.823 | 0.147 | <0.001 |
| Hospital length of stay (day) | 1.05 (1.05-1.05) | 0.050 | 0.001 | <0.001 |

### Prediction model of 30-day postoperative VTE without hospital length of stay

The prediction model for postoperative VTE using the seven predictors (excluding hospital length of stay) showed high predictive power (training cohort AUC=0.74, validation cohort AUC=0.75). Brier score was 0.008. Predictors of this model are available in **eTable I** of the **Supplement**. The calibration plot is shown in **eFigure III** of the **Supplement**.

### Application of the Risk Model

Below are example use cases for the VTE risk calculator. A VTE probability (%) can be calculated using the intercept and coefficient from multivariate logistic regression model. A web-based risk model is available for use at the bedside (http://vterisk.herokuapp.com).

1. 67 year old male, with a history of disseminated cancer, on outpatient corticosteroid treatment, no preoperative sepsis, serum albumin 2.0 g/dL, platelet count 110×10^9^/L, spine surgery: 30-day postoperative VTE risk= 6.47% (**eFigure IV** in the **Supplement**)
2. 60 year old female, no history of cancer, history of rheumatoid arthritis on corticosteroid treatment, no preoperative sepsis, serum albumin 3.5 g/dL, platelet count 130×10^9^/L, hip replacement surgery, <u>expected hospital length of stay 4 days</u>: VTE risk= 1.35%, <u>expected hospital length of stay 8 days</u>: VTE risk= 1.81%

## Discussion

Our study developed a predictive model of 30-day postoperative VTE, using a very large dataset of patients undergoing diverse types of surgery. Using eight easily available clinical and laboratory factors, this model displayed excellent predictive performance.

How could this model be used to decrease the rate of postoperative VTE? Accurate assessment of the highest risk patients and measures such as extended prophylaxis may have a significant impact on post-surgical outcomes and mortality. Despite varying VTE risk within surgery type, patient factors portend very different risks even within the same type of surgery. For example, patients with lower extremity orthopedic surgery had a wide distribution of VTE risk, from nearly 0% to over 8%, so VTE preventive measures may need to be further individualized. In patients with high VTE risk, extended use of pharmacologic therapy after discharge may decrease VTE occurrence. ACCP (American College of Chest Physicians) 2012 guidelines suggested extending thromboprophylaxis for up to 35 days for major orthopedic surgery (Grade 2B). ^13^ ACCP 2012 guidelines also recommended pharmacologic prophylaxis for 4 weeks with LMWH for patients at high risk for VTE undergoing abdominal or pelvic surgery for cancer who are not otherwise at high risk for major bleeding (Grade 1B) ^6,13^ Our study showed that 49.9% of VTE complications occurred after discharge from initial hospitalization, consistent with other studies. ^14,15^ Extended thromboprophylaxis could be more broadly applied with a better determination of an individual’s risk, which our model provides. A prospective, observational study in 2373 patients undergoing cancer surgery demonstrated that VTE was the most common cause of mortality, responsible for 46.3% of postoperative deaths. ^16^

Our study found that certain non-orthopedic surgeries, such as intestinal, vascular, and liver surgery, were associated with higher rates of DVT than orthopedic surgery. Abdominal procedures may overlap with oncologic surgery, a group of patients among which the incidence of VTE is high, with one study detecting occult postoperative DVT by surveillance duplex in 9.8% of patients despite the use of perioperative thromboprophylaxis. ^17^ For vascular surgery patients, who have a high incidence of postoperative VTE, the use of both mechanical and pharmacologic prophylaxis in the preoperative period led to a 75% reduction in postoperative VTE in one single center study.^18^ Further study is necessary to investigate whether initiation of intensive preoperative prophylaxis, and extended post-operative pharmacologic therapy would be beneficial following these highest risk procedures.

As demonstrated in the case examples, VTE risk is not fixed, but changes according to factors such as hospital length of stay. The Caprini score incorporates a large number of variables, but does not include hospital length of stay. If a patient stays in the hospital longer than expected length of stay, risk of VTE may increase, so reconsideration of VTE risk and modification of preventive measures may be necessary.

Low serum albumin was shown to be associated with an increased risk of VTE in large cohort studies and likely reflects a hyperinflammatory and/or hypercoagulable state. ^19–21^ Hypoalbuminemia was a risk factor for VTE after colorectal surgery. ^22,23^ The risk of VTE was increased among systemic glucocorticoid users, consistent with other studies. ^24,25^ As a possible explanation, one study demonstrated that glucocorticoids may increase the activity of pro-thrombotic factors. ^26^

Our study has certain strengths. First, the model was trained and validated on over 2.8 million patients across varied types of surgery, so this model is applicable to broad types of surgery compared with the modified Caprini Risk Assessment Model, which was validated in a limited number of surgery types and a significantly smaller patient population. Second, this model is derived from more recent data (2015 to 2017), when pharmacologic and mechanical thromboprophylaxis were commonly used, unlike the modified Caprini Risk Assessment Model, which was developed originally in 1991 and included only 37.2% receiving VTE prophylaxis.^8^ The use of VTE prophylaxis in surgical patients has increased significantly over the past two decades. ^27^ Third, the VTE model performance was superior or equal to other models such as Rogers (AUC 0.76), IMPROVE (AUC 0.65), and Premier (AUC 0.75). ^28 28–30 9^

The current study does have limitations. First, a history of DVTs was not included in the model as these data were not available in the ACS-NSQIP database. Second, we do not know which participants in the cohort were treated with pharmacologic preventive therapy in the perioperative period. Also, the NSQIP database does not include information on postdischarge medications, such as prolonged postdischarge VTE prophylaxis

In conclusion, this VTE risk assessment model for postoperative patients was developed and validated with a very large number of patients across a wide variety of procedures. This easy-to-use model utilizes a limited number of readily available variables and can help clinicians accurately and easily identify patients at the highest risk of VTE events, promote broader discussion and awareness between the patient and the surgeon during the perioperative period, and improve surgical morbidity and mortality.

## Data Availability

Data is available at American College of Surgeons (AC-NSQIP program)

## Acknowledgments

Dr. Woo had full access to all the data in the study and takes responsibility for the integrity of the data and the accuracy of the data analysis.

*The American College of Surgeons National Surgical Quality Improvement Program and the hospitals participating in the ACS NSQIP are the source of the data used herein; they have not verified and are not responsible for the statistical validity of the data analysis or the conclusions derived by the authors*.

## Disclosures

None

[Supplement]

**eFigure I. Study participant flow chart**

**eFigure II. Calibration plot for 30-day postoperative VTE (Model with hospital length of stay) Observed vs expected VTE risk**

**eFigure III. Calibration plot for 30-day postoperative VTE (Model without length of stay). Observed vs expected VTE risk**

**eFigure IV. Mobile web-application of VTE prediction model**

**eFigure V. Average day from operation until DVT complication according to surgery type**

**eTable I. Predictors of VTE predictors (without length of stay)**

